# Association Between the Swallowing Reflex and the Development of Aspiration Pneumonia in Patients with Dysphagia Admitted to Long-term Care Wards -A Prospective Cohort Study of 60 Days-

**DOI:** 10.1101/2020.12.20.20248528

**Authors:** Tomoya Omura, Miwa Matsuyama, Shota Nishioka, Shomu Sagawa, Masaya Seto, Mitsugu Naoe

## Abstract

**Objective:** To investigate the association between the simple swallowing provocation test (SSPT) and development of aspiration pneumonia in patients with dysphagia in long-term care (LTC) wards.

**Design:** The study design was a prospective cohort study. Subjects were followed for 60 days from admission.

**Setting:** LTC wards.

**Participants:** Study subjects were patients with dysphagia aged 65 years or older who were admitted to LTC wards between August 2018 and August 2019. In total, 39 subjects were included in the analysis (20 males, 19 females; mean age 83.8 ± 8.5 years). Subjects were divided into two groups based on SSPT results: normal swallowing reflex (SSPT normal group) and abnormal swallowing reflex (SSPT abnormal group). The covariates were age and sex, primary disease, history of cerebrovascular disease, Glasgow coma scale, body mass index, geriatric nutritional risk index, the mann assessment of swallowing ability, food intake level scale, functional independence measure, and oral health assessment tool.

**Interventions:** Not applicable.

**Main Outcome Measure:** The outcome was the incidence of aspiration pneumonia during the first 60 days of hospitalization, and the predictive factor was SSPT: 0.4 ml.

**Results:** The incidence of aspiration pneumonia was 33.3% in the SSPT normal group and 76.2% in the SSPT abnormal group. The phi coefficient was −0.43, the risk ratio was 2.29, and the 95% confidence interval (95%CI) was 1.14 to 4.58. The predictive factor for aspiration pneumonia was SSPT: 0.4 ml (95% CI: 1.57–26.03).

**Conclusions:** Our findings suggest that the SSPT provides a valid index for the development of aspiration pneumonia in older patients with dysphagia admitted to LTC wards.

## Introduction

Aspiration pneumonia is a common disease associated with older patients with dysphagia, and mortality is increasing ^1^. Aspiration pneumonia has a high recurrence rate, resulting in decreased swallowing function and diminished activities of daily living (ADL). Additionally, aspiration pneumonia leads to poor outcomes such as prolonged hospital stays and increased mortality ^2-5^. Therefore, detecting patients at risk of developing aspiration pneumonia early in hospitalization is important for subsequent prevention.

Various reports have been published on risk factors related to aspiration pneumonia ^6-9^. Among them, a review by Martino et al. reported that the risk of pneumonia was 3.17 times higher in patients with dysphagia than in patients without dysphagia, and 11.56 times higher in patients with dysphagia with aspiration ^10^. Dysphagia is an important risk factor for aspiration pneumonia ^11, 12^, and the presence of silent aspiration is known to increase the risk of aspiration pneumonia ^13, 14^. Therefore, the detection of aspiration and silent aspiration is important for predicting aspiration pneumonia.

Swallowing screening is performed to detect dysphagia and aspiration at admission, and the efficacy of some swallowing screening tests has been validated ^15-17^. However, swallowing screening is not sufficient to detect silent aspiration ^18^. The simple swallowing provocation test (SSPT) developed by Teramoto et al. is superior to other swallowing screens in evaluating the abnormal in the swallowing reflex and detecting silent aspiration ^19-21^. Additionally, it allows aspiration pneumonia to be predicted ^21^. The accuracy of predicting aspiration pneumonia is higher with SSPT than with the water swallowing test ^22^. Previous studies have found an association between the SSPT and aspiration pneumonia in patients with acute ischemic stroke and residents of a geriatric health services facility ^23, 24^. However, there have been no studies on the association between the SSPT and aspiration pneumonia in long-term care (LTC) wards that in Japan provide long-term care for older adults with severe physical and cognitive problems under the national healthcare insurance system. Patients in LTC wards are generally admitted from acute/subacute hospital wards after acute treatments, or from home due to exacerbation of their disease conditions. Japanese LTC wards are comparable to skilled nursing homes in Western countries ^25^. Pneumonia in older patients hospitalized in LTC wards is called nursing and healthcare-associated pneumonia (NHCAP, the Japanese version of healthcare-associated pneumonia) ^26^, and 63% of cases have been reported to be associated with aspiration ^27^. A previous study reported that all patients who developed NHCAP in LTC wards had dysphagia ^28^. It is presumed that patients with dysphagia admitted to LTC wards are more likely to develop aspiration pneumonia. Therefore, prediction of aspiration pneumonia is important for patients with dysphagia in LTC wards.

In this study, we aimed to clarify the association between the SSPT and aspiration pneumonia at the time of admission in patients with dysphagia admitted to LTC wards, and examined whether the SSPT can predict aspiration pneumonia.

## Methods

### Study Design

The study design was a prospective cohort study. Subjects were registered as a cohort on admission and were followed for 60 days from admission.

### Subjects

Study subjects were patients with dysphagia aged 65 years or older who were admitted to LTC wards from August 2018 to August 2019. Dysphagia was determined by the mann assessment of swallowing ability (MASA) ^29^ at admission. The total score of the MASA is 200 points, and the cutoff value is 177 points. The results of the MASA are interpreted as no abnormality (≥178), mild dysphagia (168–177), moderate dysphagia (139–167), and severe dysphagia (≤138). Subjects with 177 point or lower were registered as part of the cohort. Exclusion criteria were subjects who were discharged within 60 days, subjects with missing data, and subjects who were not assessed within 1 month of admission. This study was conducted with the approval of the Naruto Yamakami Hospital Ethics Committee (1-1-01). Informed consent was obtained in writing from the subjects. It was also obtained from an alternate in the case of subjects with communication difficulties. The analysis was conducted with consideration of protecting patients’ personal information.

The sample size was calculated based on the difference in the incidence of aspiration pneumonia between the SSPT abnormal group and the normal group in a previous study of a geriatric health services facility ^24^. The incidence of aspiration pneumonia was set at a power level of 80% and a significance level of 5%, assuming a normal group (30%) and an abnormal group (75%). It required 18 subjects in each group, making a total of 36 subjects. Exclusion criteria were subjects discharged within 60 days of admission and subjects with missing data.

### Outcome Measure

The primary outcome measure was the incidence of aspiration pneumonia during the first 60 days after hospitalization. Aspiration pneumonia was diagnosed by a physician based on the Clinical Practice Guidelines for Nursing- and Healthcare-associated Pneumonia ^26^.

### Assessment Items

The predictive factor was an SSPT with 0.4 ml water (SSPT: 0.4 ml). In previous studies, the SSPT was a two-step method based on the response to 0.4ml water with an additional test with 2 ml ^21^. However, in this study, the response to 0.4ml water alone was used to simplify the examination and the result determination. The SSPT was conducted as described by Teramoto et al. ^20, 21^. SSPT measurements were performed with the patient in a supine position. A 5Fr feeding catheter^a^ (outside diameter 1.7 mm, length 40 cm) was inserted nasally for about 13 cm and placed in the oropharynx. The swallowing reflex was induced by bolus injection of 0.4 ml of distilled water. The water bolus injection was administered near the end of expiration. The latent time from the beginning of the water bolus injection to the onset of swallowing (i.e., laryngeal elevation) was measured with a stopwatch. The swallowing reflex was confirmed by visual inspection of laryngeal elevation. The cutoff was set to 3 seconds. A swallowing reflex observed within 3 seconds was classified as normal, and no reflex within 3 seconds was classified as abnormal. A total of three trials were performed, and the average of two short latencies was used. The covariates were age and sex, primary disease, history of cerebrovascular disease, Glasgow coma scale (GCS), MASA, food intake level scale (FILS) ^30^, body mass index (BMI), geriatric nutritional risk index (GNRI, nutrition index of older adults) ^31^, functional independence measure (FIM, ADL index), oral health assessment tool (OHAT, index of oral condition) ^32^. All (covariates) data were assessed within 1 month of admission.

### Statistical Analysis

Subjects were divided into two groups: those with a normal swallowing reflex (SSPT normal group) and those with an abnormal swallowing reflex (SSPT abnormal group) based on the SSPT results. Descriptive statistics were performed after data acquisition, and we performed comparisons between the two groups for each assessment item. Multivariate analysis was used to extract predictive factors associated with aspiration pneumonia. Mann–Whitney U tests, Fisher’s exact test, and chi-square tests were used for comparisons between the two groups. Chi-square tests were also used to calculate the phi coefficient, risk ratio, and 95% confidence interval (95%CI) between SSPT: 0.4 ml and aspiration pneumonia. Predictive factors were extracted by logistic regression analysis, with aspiration pneumonia as the objective variable. SSPT: 0.4 ml and assessment items that showed a significant difference between the two groups were used as explanatory variables.

We also calculated the sensitivity, specificity, and positive predictive value of SSPT: 0.4 ml to confirm the prediction accuracy of aspiration pneumonia. We used IBM SPSS Statistics version 25^b^ for statistical analyses, with the significance level set at < 5%.

## Results

We enrolled 54 consecutive patients with dysphagia who were hospitalized during the study period. Subjects excluded were those who were discharged within 60 days of admission (n = 7), those with missing data (n = 3), and those who were not assessed within 1 month of admission (n = 5). In total, 39 subjects were included in the analysis (20 males, 19 females; mean age 83.8 ± 8.5 years). Eighteen cases were allocated to the SSPT normal group, and 21 cases were allocated to the SSPT abnormal group.

Twenty-two subjects (56.4%) developed aspiration pneumonia within 60 days of hospitalization. There was a significant difference in the incidence of aspiration pneumonia between the two groups. The incidence rate was 33.3% (6 cases) in the SSPT normal group and 76.2% (16 cases) in the SSPT abnormal group (Fig. 1).

**Fig.1.**
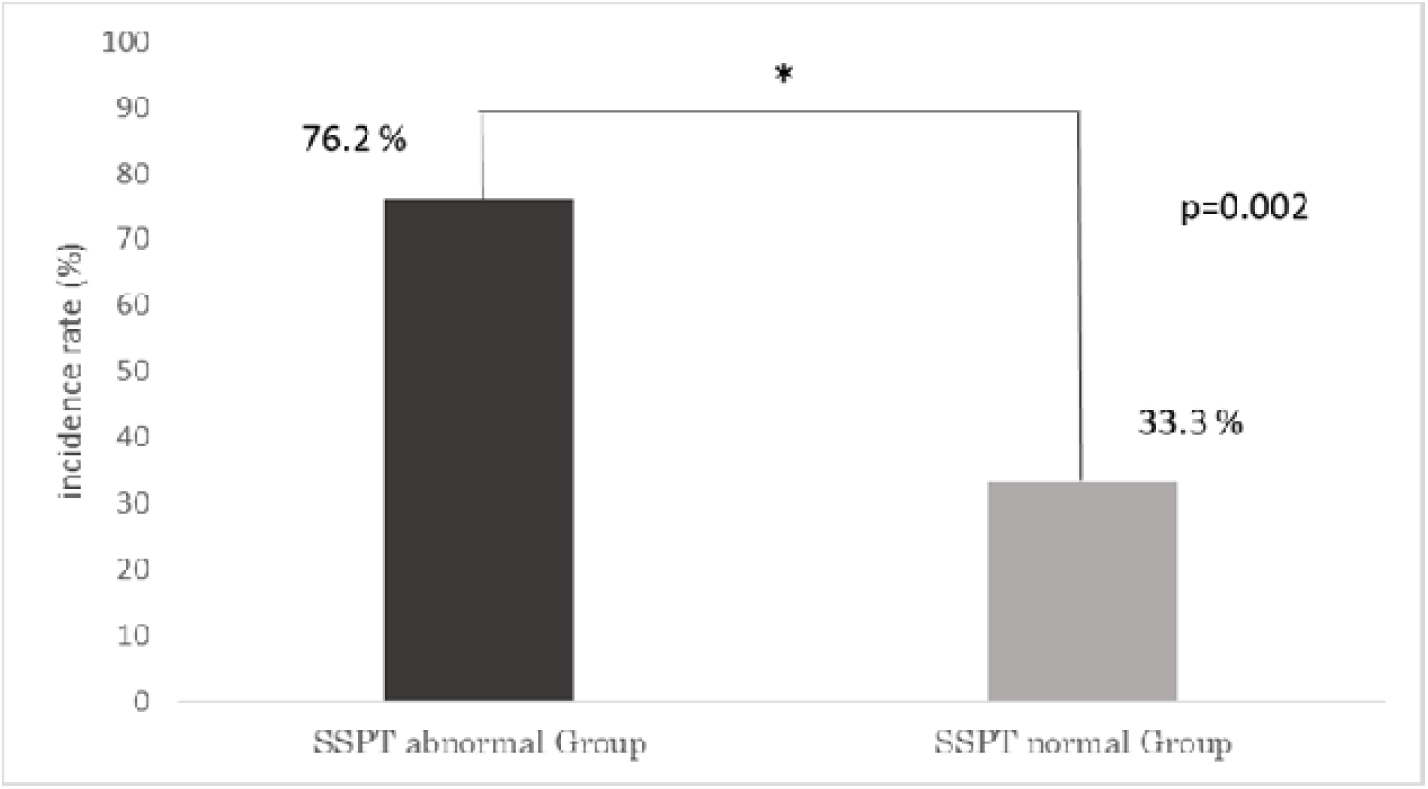
Incidence of aspiration pneumonia.

Comparison between the two groups revealed significant differences in the assessment items of age, sex, and primary disease (disuse syndrome was more common in the SSPT normal group) (Table 1).

**Table 1.** Subjects’ basic attributes

|  | Total<br>(n = 39) | SSPT abnormal group<br>(n = 21) | SSPT normal group<br>(n = 18) | p-value |
| --- | --- | --- | --- | --- |
| Age (years), mean ± SD | 83.8 ± 8.5 | 81.1 ± 8.6 | 86.9 ± 7.3 | 0.032 |
| Sex (n), male/female | 20/19 | 15/6 | 5/13 | 0.007 |
| Primary disease |  |  |  | 0.018 |
| Cerebrovascular disease, n (%) | 15 (38%) | 10 (48%) | 5 (28%) | 0.204 |
| Disuse syndrome, n (%) | 17 (44%) | 5 (24%) | 12 (67%) | 0.007 |
| Parkinson's disease, n (%) | 2 (5%) | 2 (10%) | 0 (0%) | 0.490 |
| Cancer, n (%) | 1 (3%) | 0 (0%) | 1 (6%) | 0.462 |
| Chronic obstructive pulmonary<br>disease, n (%) | 2 (5%) | 2 (10%) | 0 (0%) | 0.490 |
| Orthopedic disease, n (%) | 2 (5%) | 2 (10%) | 0 (0%) | 0.490 |
| History of cerebrovascular disease<br>Yes/No, n | 22/17 | 11/10 | 11/7 | 0.584 |
| GCS, median (range) | 13 (11–14) | 13 (11–4) | 12 (11–14) | 0.876 |
| MASA, median (range) | 140 (94–151) | 134 (92–147) | 141 (110–151) | 0.499 |
| FILS, median (range) | 2 (2–7) | 2 (2–7) | 3 (2–6) | 0.770 |
| BMI, median (range) | 18.0 (16.1–19.5) | 18.1 (16.8–19.3) | 18.0 (16.0–21.6) | 0.833 |
| GNRI, median (range) | 76.5 (66.5–83.5) | 79.2 (67.6–92.3) | 70.5 (65.5–85.7) | 0.220 |
| FIM (score), median (range) | 23 (18–32) | 23 (18–35) | 20 (18–30) | 0.852 |
| OHAT (score), median (range) | 6 (4–8) | 6 (4–8) | 6 (3–7) | 0.487 |

The phi coefficient was −0.43, the risk ratio was 2.29, and the 95%CI was 1.14 to 4.58 for the SSPT abnormal group (Table 2).

**Table 2.**
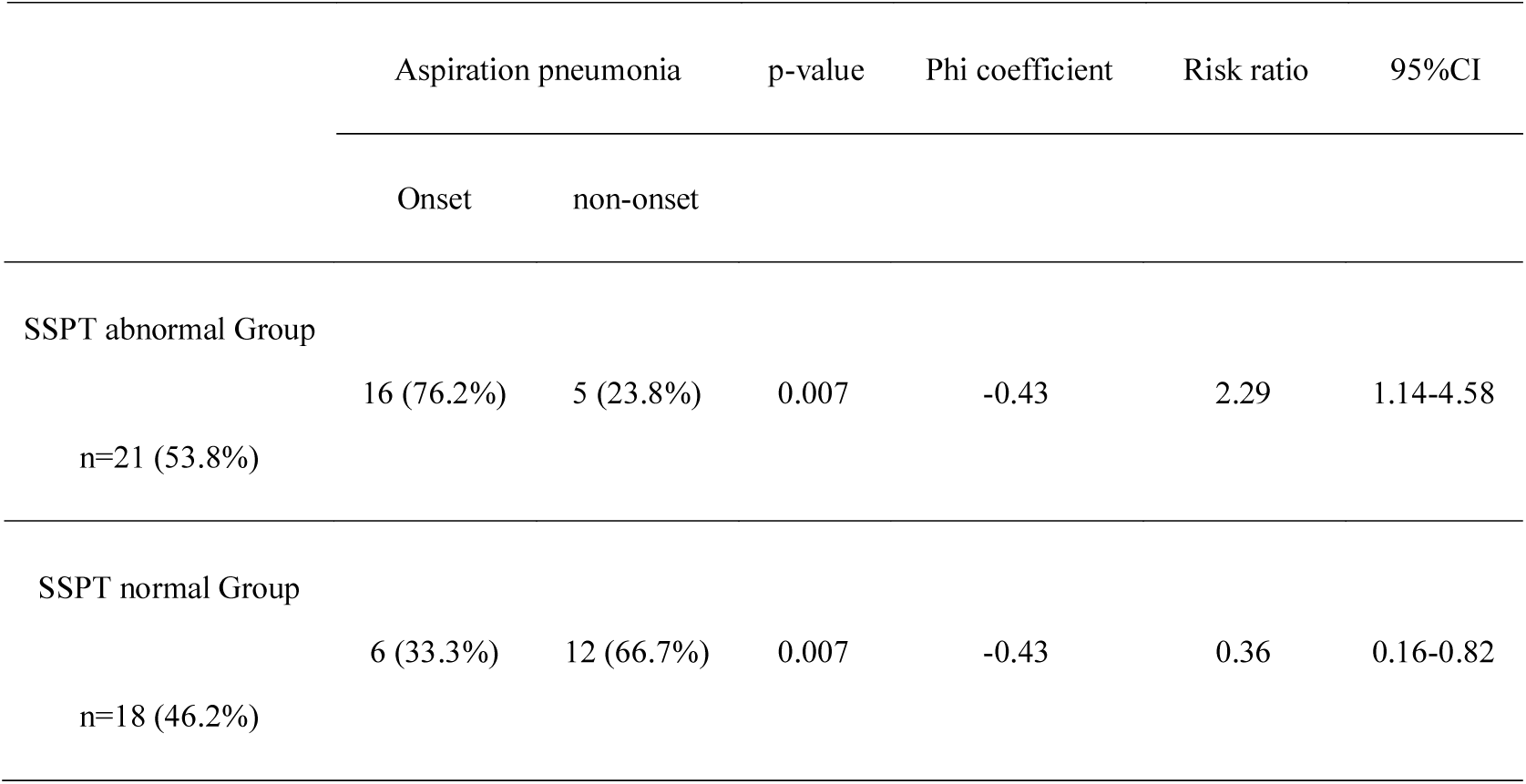
Phi coefficient and risk ratios for aspiration pneumonia

Logistic regression analysis showed that SSPT: 0.4 ml was a predictive factor associated with aspiration pneumonia. The odds ratio was 6.40 and the 95%CI was 1.57–26.03 (Table 3).

**Table 3.** Predictive factors associated with aspiration pneumonia

|  | Regression coefficient | p-value | Odds ratio | 95%CI |
| --- | --- | --- | --- | --- |
| SSPT abnormal | 1.86 | 0.01 | 6.40 | 1.57 - 26.03 |

Objective variable: Aspiration pneumonia
Explanatory variables: SSPT: 0.4 ml, Age, Sex, Primary disease

The prediction accuracy of SSPT: 0.4 ml for aspiration pneumonia was 73% sensitivity, 71% specificity, and 76% positive predictive value (Table 4).

**Table 4.** Predictive accuracy of aspiration pneumonia

|  | Sensibility | Specificity | Positive predictive value |
| --- | --- | --- | --- |
| Aspiration pneumonia | 73 | 71 | 76 |

## Discussion

The SSPT measures the swallowing reflex with a small amount of water injected through a 5Fr feeding catheter inserted nasally 13 cm into the oropharynx ^20, 21^. This test can detect the risk of silent aspiration ^19, 20^ and can predict aspiration pneumonia ^21, 23, 24^. The SSPT has the advantage of being able to be performed on patients with dementia or communication disorders ^33^. However, the disadvantage of SSPT is that it only evaluates the pharyngeal swallowing reflex. In this study, we investigated the association between the swallowing reflex in the SSPT and the development of aspiration pneumonia in older patients with dysphagia after the acute phase by means of a prospective cohort study. The results of this study showed that the development of aspiration pneumonia was 2.3 times higher in patients with an abnormal swallowing reflex in the SSPT than in those a normal swallowing reflex. We hypothesized that the SSPT could be a screening test to predict the development of aspiration pneumonia in older patients with dysphagia after the acute phase.

Several studies have investigated the association between SSPT and aspiration pneumonia in patients with acute ischemic stroke and residents of a geriatric health services facility ^23, 24^. In contrast, we examined older patients with dysphagia after the acute phase with the SSPT and, while our findings supported those of previous studies, we found that the incidence of aspiration pneumonia was higher than that of patients with acute ischemic stroke (14%) ^23^. It is presumed that the subjects of this study were patients with many potential risks for aspiration pneumonia at baseline. The subjects of the previous study were patients with acute ischemic stroke, with a median age of 73 years, and 54% of the patients were discharged home ^23^. The subjects in this study had a high average age of 83.8 years, with median scores of MASA: 140, FILS: 2, FIM: 23, and GNRI: 76.5. All had poor swallowing function, ADL, and nutritional status and required ongoing medical care. Age, dysphagia, low ADL, and malnutrition have been reported as risk factors for aspiration pneumonia ^28, 34, 35^. The subjects in this study were about 10 years older than those in the previous study and had a number of risk factors for aspiration pneumonia. Furthermore, the previous study had sensitivity and specificity of 66.7% and 83.5%, compared with 73% and 71% in this study ^23^. Although aspiration does not necessarily lead directly to the development of aspiration pneumonia ^10^, the incidence of aspiration pneumonia increases as the number of risk factors associated with aspiration pneumonia increases ^33^. This suggests that an abnormal swallowing reflex was more likely to lead to the development of aspiration pneumonia in the subjects of this study who had many risk factors for aspiration pneumonia. Therefore, the sensitivity may have been higher than in previous studies. Hence, in patients at high risk for aspiration pneumonia, the abnormal swallowing reflex may increase the incidence of aspiration pneumonia.

In contrast with the previous studies that were retrospective ^23, 24^, our study was a prospective cohort study with a total of 39 cases (18 in the normal group and 21 in the abnormal group). The phi coefficient was −0.43, indicating that the effect size was sufficient. The width of the 95%CI for the risk ratio was narrow at 3.44, suggesting that the sample size was appropriate.

About 80% of patients admitted to LTC wards are over 75 years old ^36^, and 72.1% of patients have low ADL levels and need medical care ^25^. The demographics of the subjects in our study were similar to this, with an average age of 83.3 years and low ADL levels. Therefore, it is speculated that many LTC wards have similar characteristics to this study. The findings in this study may predict the development of aspiration pneumonia in older patients with dysphagia after the acute phase.

### Study Limitations

This study has some limitations. First, the SSPT may not adequately detect the risk of aspiration in patients with a sole or predominant impairment in the oral phase of swallowing and a relatively intact pharyngeal phase ^37^. In fact, it was inferred from the findings that 33.3% of the SSPT normal group developed aspiration pneumonia. Second, this study investigated subjects in a single institution, raising an issue with external validity. Further studies should include data from multiple institutions in combination with other screening methods to further improve the prediction accuracy.

## Conclusions

This is the first prospective cohort study to examine the association between the swallowing reflex and the development of aspiration pneumonia in older patients with dysphagia (mean age 83.8 years) in LTC wards. The findings of this study suggest that the abnormal of a swallowing reflex in the SSPT with 0.4 ml water was associated with the development of aspiration pneumonia, and the abnormal of a swallowing reflex could have 2.3 times the risk of developing aspiration pneumonia when compared with those the normal reflex. We concluded that SSPT could be a predictive factor for aspiration pneumonia in older LTC inpatients.

## Data Availability

The data that support the findings of this study are available from the corresponding author, Omura, T., upon reasonable request.

## List of abbreviations

SSPT: the simple swallowing provocation test
LTC: long-term care
ADL: activities of daily living
NHCAP: nursing and healthcare-associated pneumonia
MASA: the mann assessment of swallowing ability
GCS: glasgow coma scale
FILS: food intake level scale
BMI: body mass index
GNRI: geriatric nutritional risk index
FIM: functional independence measure
OHAT: oral health assessment tool
CI: confidence interval

## Acknowledgments

The authors thank all members of the Naruto-Yamakami Hospital for their help with this research. We also thank Helen Jeays, BDSc AE, from Edanz Group (https://en-author-services.edanzgroup.com/ac) for editing a draft of this manuscript.

## Suppliers

a. 5Fr atom feeding catheter; Atom Medical Corporation, 3-18-15, Hongo, Bunkyo-ku, Tokyo to, 113-0033, Japan
b. IBM SPSS Statistics version 25; IBM Japan, 19-21, Nihombashihakozakicho, Chuo-ku, Tokyo to, 103-8510, Japan

## Notes

### Competing Interest Statement

The authors have declared no competing interest.

### Funding Statement

This research did not receive any specific grant from funding agencies in the public, commercial, or not-for-profit sectors.

### Author Declarations

This study was conducted with the approval of the Naruto Yamakami Hospital Ethics Committee (1-1-01).

